# ADCC-activating antibodies correlate with protection against congenital human cytomegalovirus infection

**DOI:** 10.1101/2023.03.15.23287332

**Authors:** Eleanor C. Semmes, Itzayana G. Miller, Nicole Rodgers, Caroline T. Phan, Jillian H. Hurst, Kyle M. Walsh, Richard J. Stanton, Justin Pollara, Sallie R. Permar

**Affiliations:** Medical Scientist Training Program, Department of Molecular Genetics and Microbiology, Duke University, Durham, NC, USA; Duke Human Vaccine Institute, Duke University, Durham, NC, USA; Department of Surgery, Duke University School of Medicine, Durham, NC 27710, USA; Department of Pediatrics, Duke University, Durham, NC, USA; Department of Neurosurgery, Duke University, Durham, NC, USA; Division of Infection and Immunology, School of Medicine, Cardiff University, Cardiff, UK; Department of Pediatrics, Weill Cornell Medicine, New York City, NY, USA

## Abstract

Human cytomegalovirus (HCMV) is the most common vertically transmitted infection worldwide, yet there are no licensed vaccines or therapeutics to prevent congenital HCMV (cCMV) infection. Emerging evidence from studies of natural infection and HCMV vaccine trials indicates that antibody Fc effector functions may defend against HCMV infection. We previously reported that antibody-dependent cellular phagocytosis (ADCP) and IgG activation of FcγRI/FcγRII were associated with reduced risk of cCMV transmission, leading us to hypothesize that other Fc-mediated antibody functions may also contribute to protection. In this same cohort of HCMV transmitting (n = 41) and non-transmitting (n = 40) mother-infant dyads, we found that higher maternal sera antibody-dependent cellular cytotoxicity (ADCC) activation was also associated with decreased risk of cCMV infection. We determined that NK cell-mediated ADCC responses correlated strongly with anti-HCMV IgG FcγRIII/CD16 activation and IgG binding to the HCMV immunoevasin protein UL16. Notably, anti-UL16 IgG binding and engagement of FcγRIII/CD16 were higher in non-transmitting versus transmitting dyads and interacted significantly with ADCC responses. These findings indicate that ADCC-activating antibodies against novel targets such as UL16 may represent an important protective maternal immune response against cCMV infection, which can guide future HCMV correlates studies and vaccine development.

## INTRODUCTION

Human cytomegalovirus (HCMV) is the most common vertically transmitted infection worldwide and has been associated with stillbirth, neurodevelopmental impairment, sensorineural hearing loss, and childhood leukemia (1, 2). Over 80% of reproductive age women worldwide are HCMV seropositive and congenital transmission can occur following primary or non-primary HCMV infection, which may include reinfection with a new strain or reactivation from viral latency (3). Despite these disease risks and the ubiquity of congenital HCMV (cCMV) infection, we lack effective therapeutics and vaccines to prevent HCMV transmission. Neutralizing antibodies against HCMV envelope glycoproteins (e.g., gB and pentamer complex) and T cell responses have been the main targets in vaccine development to-date, but these vaccines have achieved only limited to moderate efficacy (4). Moreover, several studies have found that maternal neutralizing antibody titers do not correlate with reduced risk of cCMV infection (5, 6). We also recently reported that neutralizing antibody titers against multiple HCMV strains and cell types were higher magnitude in HCMV transmitting pregnancies and not associated with protection (7). Maternal administration of HCMV hyperimmunoglobulin (HCMV-HIG), a polyclonal preparation of IgG from HCMV seropositive donors, following primary infection during pregnancy also failed to prevent congenital transmission in two randomized clinical trials (8, 9), despite suggested efficacy in observational studies (10-12). Thus, an improved understanding of the maternal antibody responses that protect against cCMV infection is urgently needed to guide the development of novel vaccines and immunotherapeutics (13, 14).

Antibodies can mediate polyfunctional responses such as viral neutralization through the Fab region that binds antigen and non-neutralizing effector functions through the constant Fc region that binds Fc receptors (FcRs) on innate immune cells. Emerging evidence indicates that anti-viral IgG effector functions mediated by interactions between the IgG Fc region and FcγRs are a previously underappreciated component of anti-HCMV immunity (15, 16). In our recent maternal-infant dyad study reporting that neutralizing antibody titers were not correlated with protection, we found that greater Fc-mediated antibody responses were associated with decreased risk of HCMV transmission (7). Specifically, higher maternal sera anti-HCMV IgG engagement of FcγRI/FcγRIIa and activation of antibody-dependent cellular phagocytosis (ADCP) were associated with protection against cCMV infection. Therefore, we posited that additional antibody Fc effector functions may be important.

Antibody-dependent cellular cytotoxicity (ADCC) is an anti-viral Fc effector function that has been underexplored in cCMV infection to-date. Natural killer (NK) cells can eliminate virally-infected cells via ADCC, which is mediated by FcγRIII/CD16 expressed on the NK cell surface, or through direct cytotoxic killing. HCMV has evolved many mechanisms to evade antibody-dependent and -independent NK cell killing by encoding “immunoevasins” that interfere with the host immune response (17, 18). Specific immune evasion strategies employed by HCMV include viral FcγR decoys that sequester IgG to prevent FcγRIII activation (19, 20) and viral proteins that modulate NK cell cytotoxicity (18, 21). Nevertheless, NK cell-mediated ADCC can inhibit cell-to-cell spread of HCMV in multiple cell types (22, 23), even with strains that express these viral immunoevasins (24). Paradoxically, some NK cell immunoevasins (e.g., viral FcγR gp34, UL16, and UL141) have even been identified as targets of ADCC-activating IgG (24). Despite evidence that ADCC mediated by certain IgG specificities can overcome viral immune evasion, whether ADCC or anti-immunoevasion antibodies help limit HCMV transmission in utero is unknown.

In this study, we hypothesized that greater ADCC responses and FcγRIII/CD16 engagement by maternal antibodies would be correlated with decreased risk of cCMV infection. To investigate this hypothesis, we quantified ADCC responses and FcγRIII activation in maternal and cord blood sera from HCMV transmitting and non-transmitting pregnancies (7). We then explored whether HCMV-specific IgG directed against different antigens including envelope glycoproteins, tegument proteins, and immunoevasins might contribute to protective ADCC responses. Our findings support an accumulating body of evidence that Fc-mediated immunity and IgG directed against non-canonical HCMV antigens are important for anti-viral control and should be considered in future correlates and vaccine studies.

## RESULTS

### Overview of mother-infant cohort

To determine whether ADCC-mediating antibodies may protect against cCMV, we compared antibody responses in maternal and cord blood sera from HCMV seropositive transmitting (n = 41) and non-transmitting (n = 40) mother-infant dyads that we previously identified as donors to a large US-based public cord blood bank (**Supplementary Figure 1**) (7). Cases of cCMV infection were identified based on the detection of HCMV DNAemia in the cord blood plasma and dyads were matched on infant sex, race, maternal age, and delivery year. Cord blood donors were screened for clinical signs of (a) neonatal sepsis, (b) congenital infection (petechial rash, hepatosplenomegaly, thrombocytopenia), and (c) congenital abnormalities at birth, and only term, healthy, uncomplicated births were included.

Demographic and clinical characteristics were comparable between groups (**Table 1**) (7), but HCMV serologies differed between transmitting and non-transmitting dyads. Median HCMV IgG relative avidity index (RAI) scores were lower in transmitting (median = 67.7%) versus non-transmitting (median = 74.8%) pregnancies, and 19.5% of transmitting cases were classified as low/intermediate avidity (<60%), whereas only 2.5% of non-transmitting pregnancies had low/intermediate RAI scores (**Table 1**). Additionally, 26.8% of transmitting pregnancies had HCMV-specific IgM in maternal sera versus only 5% of non-transmitting pregnancies (**Table 1**). While it is not possible to define primary infection, reinfection, or reactivation based on HCMV serologies at a single timepoint (25, 26), these data imply that the timing of maternal HCMV exposure may have differed between groups.

**Table 1.** Overview of case-control cord blood donor mother-infant cohort.

|  | **HCMV transmitting (n=41)** | **HCMV non-transmitting (n=40)** |
| --- | --- | --- |
| **Infant sex, n (%)** |  |  |
| **Female** | 17 (41.5%) | 16 (40.0%) |
| **Male** | 24 (58.5%) | 24 (60.0%) |
| **Infant race/ethnicity, n (%)** |  |  |
| **White** | 26 (63.4%) | 24 (60.0%) |
| **Black or African American** | 8 (19.5%) | 8 (20.0%) |
| **Hispanic or Latino** | 2 (4.9%) | 2 (5.0%) |
| **Other** | 5 (12.2%) | 6 (15.0%) |
| **Maternal age (years), median [IQR]** | 27 [23, 31] | 28 [24, 33] |
| **Gestational age (months), median [IQR]** | 39.0 [39.0, 40.0] | 39.0 [38.0, 40.0] |
| **Delivery year, median [range]** | 2013 [2010, 2015] | 2012 [2008, 2017] |
| **Delivery type, n (%)** |  |  |
| **Vaginal** | 18 (43.9%) | 24 (60.0%) |
| **Cesarean section** | 23 (56.1%) | 16 (40.0%) |
| **Maternal HCMV IgG avidity score<sup>a</sup>, median [IQR]** | 67.7 [63.8-70.9] | 74.8 [71.9-78.3] |
| **Maternal HCMV IgG avidity score<sup>a</sup>, n (%)** |  |  |
| **Low/intermediate (< 60%)** | 8 (19.5%) | 1 (2.5%) |
| **High (≥ 60%)** | 33 (80.5%) | 39 (97.5%) |
| **Maternal HCMV IgM seropositivity, n (%)** |  |  |
| **Seropositive** | 11 (26.8%) | 2 (5.0%) |
| **Seronegative** | 30 (73.2%) | 38 (95.0%) |
| **Maternal sera HCMV DNAemia, n (%)** |  |  |
| **Positive** | 11 (26.8%) | 15 (37.5%) |
| **Negative** | 30 (73.2%) | 25 (62.5%) |
| **Maternal sera HCMV viral copies, median [range]<sup>b</sup>** | 346 [256-1052] | 365 [260-719] |
| **Cord blood sera HCMV viral copies, median [range]<sup>b</sup>** | 727 [137-18,100] | ND |

ND = not detected
<sup>a</sup> Maternal HCMV IgG relative avidity index (RAI) score was calculated as the mean HCMV-specific IgG RAI measured against 3 HCMV strains including TB40E, AD169r, and Toledo virus. RAI scores <60% were considered low/intermediate avidity whereas scores ≥60% were considered high avidity based on numerous previous publications (Prince et al.).
<sup>b</sup> Maternal HCMV viral copies listed in viral copies/mL, lower limit of detection = 250 copies/mL
<sup>c</sup> Cord blood HCMV viral copies detected listed in IU/mL, lower limit of detection = 137 copies/mL
Transmitting and non-transmitting dyads were matched on maternal age (+/- 3 years), infant race, sex, and delivery year (+/- 3 years)

### HCMV-specific ADCC and FcγRIII/CD16 activating antibodies are higher in non-transmitting versus transmitting mother-infant dyads

Using NK cell degranulation (i.e., % CD107a positive NK cells) against HCMV-infected fibroblasts to quantify ADCC (**Supplementary Figure 2A-B, Figure 1A**), we found that antibodies in sera from non-transmitting dyads mediated significantly greater ADCC compared to transmitting dyads (**Figure 1B, Supplementary Tables 1-2**). ADCC responses in paired cord blood and maternal sera were strongly correlated, yet NK cell degranulation was significantly lower in cord blood versus maternal sera (**Figure 1B**). Few dyads had placental IgG transfer ratios >1.0 or 100% (**Figure 1B**), suggesting that the transfer efficiency of ADCC-activating IgG into fetal circulation was poor. In our univariate regression analysis, higher ADCC activation in maternal (OR = 0.86, p = 0.016) and cord blood sera (OR = 0.79, p = 0.005) were associated with reduced risk of cCMV (**Tables 2-3**). Moreover, ADCC-activating antibodies were associated with a greater magnitude reduction in transmission risk compared to our previously identified immune correlate of ADCP-activating antibodies (OR = 0.94, p = 0.008) (7) (**Tables 2-3**). To assess if differences in maternal HCMV exposure may be confounding these results, we compared ADCC in dyads stratified by RAI score or HCMV-specific IgM status and found that ADCC activation was lower in dyads with low/intermediate (median = 5.1%) versus high (median = 9.5%, p = 0.001) RAI scores (**Supplementary Tables 3-4**). In a sensitivity analysis excluding dyads with low/intermediate RAI scores, ADCC remained significantly associated with protection against transmission in cord blood sera (OR = 0.81, p = 0.017) with a trend towards significance in maternal sera (OR = 0.89, p = 0.065, **Supplementary Table 5**).

**Figure 1.**
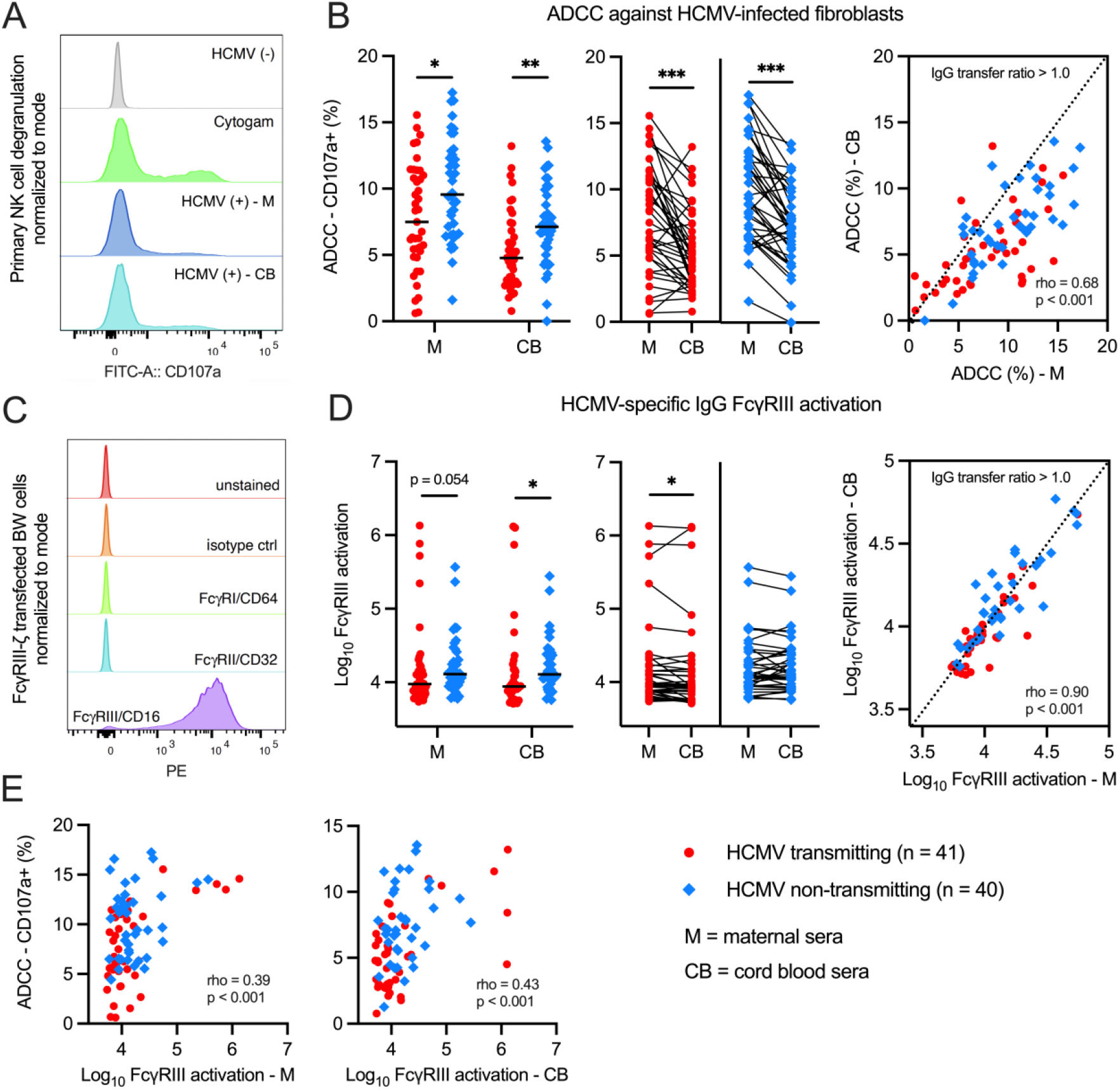
HCMV-specific ADCC and FcγRIII/CD16 activating antibodies in HCMV transmitting versus non-transmitting mother-infant dyads. HCMV-specific ADCC and FcγRIII IgG activation was measured using maternal (M) and cord blood (CB) sera from HCMV transmitting (red circles, n = 41) and non-transmitting (blue diamonds, n = 40) mother-infant dyads. Antibody responses were compared between and within dyads. (A) NK cell degranulation (% CD107a positive NK cells; gating strategy in Supplementary Figure 2) was quantified as a read-out of ADCC using a flow-based assay. Primary NK cells were isolated from PBMCs by negative selection with magnetic beads prior to co-incubation with HCMV-infected and mock-infected cells. Cytogam (light green), HCMV seropositive (light and dark blue), and HCMV seronegative (grey) sera samples were included as controls. (B) HCMV-specific antibody ADCC responses in transmitting and non-transmitting dyads. (C) Flow cytometry of FcγR-CD3ζ BW cells showing unstained (red), isotype control (orange), anti-FcγRI/CD64 (light green), anti-FcγRII/CD32 (blue), and anti-FcγRIII/CD16 (purple) PE-conjugated antibody staining. (D) Anti-HCMV IgG FcγRIII activation in transmitting and non-transmitting dyads. (E) Scatterplots showing Spearman correlations between HCMV-specific ADCC and FcγRIII IgG activation. IgG transfer ratio equals paired cord blood/maternal sera responses. Horizontal black bars denote median. FDR-corrected *P* values for Mann-Whitney U test or Wilcoxon signed-rank test. * *P* < 0.05, \*\**P* < 0.01, \*\*\**P* < 0.001.

**Table 2.** Univariate logistic regression and interaction analysis of maternal sera antibody responses and risk of congenital HCMV transmission.

| Antibody variable | Univariate |  |  | HCMV ADCC Interaction |  |  |
| --- | --- | --- | --- | --- | --- | --- |
|  | OR<sup>a</sup> | 95% CI | p value | R<sup>b</sup> | 95% CI | p value |
| **HCMV ADCC** | 0.86 | 0.77 - 0.97 | **0.016** | - | - | - |
| **HCMV ADCP<sup>c</sup>** | 0.94 | 0.90 - 0.98 | **0.008** | 0.99 | 0.98 - 1.00 | 0.079 |
| **HCMV IgG FcγRIII activation** | 0.93 | 0.63 - 1.37 | 0.700 | 1.30 | 1.08 - 1.57 | **0.005** |
| **UL141 IgG binding level** | 2.53 | 1.31 - 4.89 | **0.001** | 0.95 | 0.81 - 1.13 | 0.586 |
| **UL141 IgG FcγRIII V158** | 0.99 | 0.67 - 1.46 | 0.962 | 1.09 | 0.98 - 1.23 | 0.113 |
| **UL141 IgG FcγRIII F158** | 0.97 | 0.66 - 1.43 | 0.899 | 1.05 | 0.93 - 1.19 | 0.439 |
| **UL16 IgG binding level** | 0.79 | 0.60 - 1.05 | 0.101 | 1.12 | 1.02 - 1.23 | **0.023** |
| **UL16 IgG FcγRIII V158** | 0.70 | 0.58 - 0.86 | **0.001** | 1.06 | 0.99 - 1.12 | 0.071 |
| **UL16 IgG FcγRIII F158** | 0.75 | 0.63 - 0.88 | **0.001** | 1.06 | 1.01 - 1.12 | **0.025** |
| **UL16 IgG FcγRIII activation** | 0.82 | 0.70 - 0.96 | **0.012** | 1.05 | 1.00 - 1.10 | **0.037** |

<sup>a</sup> OR < 1.0 is associated with decreased risk and OR > 1.0 is associated with increased risk of congenital HCMV transmission.
<sup>b</sup> R = ratio of odds ratios for the interaction term between HCMV ADCC (measured as NK cell degranulation) and HCMV-specific antibody responses.
<sup>c</sup> Antibody-dependent cellular phagocytosis (ADCP) was measured previously in Semmes et al. 2022.
Bold indicates statistical significance (p < 0.05)

**Table 3.** Univariate logistic regression and interaction analysis of cord blood sera antibody responses and risk of congenital HCMV transmission.

| Antibody variable | Univariate |  |  | HCMV ADCC Interaction |  |  |
| --- | --- | --- | --- | --- | --- | --- |
|  | OR<sup>a</sup> | 95% CI | p value | R<sup>b</sup> | 95% CI | p value |
| **HCMV ADCC** | 0.79 | 0.68 - 0.93 | **0.005** | - | - | - |
| HCMV ADCP<sup>c</sup> | 1.00 | 0.96 - 1.04 | 0.944 | 1.01 | 1.00 - 1.02 | 0.231 |
| **HCMV IgG FcγRIII activation** | 0.93 | 0.65 - 1.33 | 0.696 | 1.21 | 1.02 - 1.43 | **0.026** |
| **UL141 IgG binding level** | 1.97 | 1.12 - 3.47 | **0.018** | 1.05 | 0.89 - 1.23 | 0.567 |
| **UL141 IgG FcγRIII V158** | 1.12 | 0.77 - 1.64 | 0.558 | 1.21 | 0.99 - 1.47 | 0.057 |
| **UL141 IgG FcγRIII F158** | 1.22 | 0.76 - 1.99 | 0.406 | 1.08 | 0.90 - 1.30 | 0.381 |
| **UL16 IgG binding level** | 0.75 | 0.55 - 1.00 | 0.051 | 1.10 | 0.99 - 1.24 | 0.073 |
| **UL16 IgG FcγRIII V158** | 0.69 | 0.56 - 0.85 | **0.001** | 1.04 | 0.96 - 1.12 | 0.323 |
| **UL16 IgG FcγRIII F158** | 0.73 | 0.61 - 0.87 | **0.001** | 1.03 | 0.97 - 1.10 | 0.294 |
| **UL16 IgG FcγRIII activation** | 0.95 | 0.83 - 1.08 | 0.472 | 1.03 | 0.97 - 1.08 | 0.319 |

<sup>a</sup> OR < 1.0 is associated with decreased risk and OR > 1.0 is associated with increased risk of congenital HCMV transmission.
<sup>b</sup> R = ratio of odds ratios for the interaction term between HCMV ADCC (measured as NK cell degranulation) and HCMV-specific antibody responses.
<sup>c</sup> Antibody-dependent cellular phagocytosis (ADCP) was measured previously in Semmes et al. 2022.
Bold indicates statistical significance (p < 0.05)

Since ADCC is primarily mediated by FcγRIII/CD16 on NK cells binding to the IgG Fc region, we hypothesized that HCMV-specific IgG in non-transmitting dyads may have enhanced FcγRIII engagement compared to transmitting dyads. To explore this hypothesis, we quantified anti-HCMV IgG activation of FcγRIII using reporter cells expressing chimeric human FcγRIII fused to a mouse CD3ζ signaling domain (**Figure 1C**). Non-transmitting dyads had significantly higher HCMV-specific IgG FcγRIII activation in cord blood (p = 0.008) with a trend towards significance in maternal sera (p = 0.054) (**Figure 1D, Supplementary Tables 1-2**). FcγRIII activation was lower in cord blood (median = 8791 μg/mL) versus maternal sera (median = 9481 μg/mL, p = 0.032) in transmitting but not non-transmitting dyads (**Figure 1D**), suggesting that infected infants may receive less FcγRIII-activating IgG via placental transfer. HCMV-specific IgG FcγRIII activation correlated with ADCC (**Figure 1E**) but was not independently associated with transmission risk in our univariate analysis (**Tables 2-3**). Nevertheless, our interaction analysis demonstrated that the association between ADCC and reduced risk of cCMV infection was stronger in dyads with greater FcyRIII activation (**Tables 2-3**), suggesting that better FcyRIII engagement contributed to protective ADCC responses.

### HCMV-specific ADCC and FcγRIII/CD16 activation correlate with anti-UL16 and anti-UL141 IgG

Having identified that ADCC-activating antibodies were associated with protection against cCMV, we aimed to define which antibodies may be contributing to this response. We hypothesized that certain IgG specificities against viral immunoevasins (e.g., UL16- and UL141-specific IgG) may activate greater ADCC responses in non-transmitting dyads (24). We also explored IgG responses against HCMV envelope glycoproteins (e.g., gB, gH/gL, gH/gL/gO, pentamer complex) and tegument proteins (e.g., pp52, pp28, pp150) since whether they activate ADCC has been poorly characterized.

We quantified IgG binding levels against 9 HCMV antigens and measured how well these antibodies bound FcγRIII. Since FcγRIII binding is influenced by IgG Fc region characteristics as well as FcγRIII genetic polymorphisms, we included binding to both the high-affinity (V158) and low-affinity (F158) FcγRIII variants. Hierarchical clustering identified 3 distinct clusters of antibody responses (**Figure 2**). HCMV-specific ADCC (i.e., % CD107a positive NK cells), FcγRIII activation, anti-UL16, and anti-UL141 IgG responses correlated together in cluster 1, whereas cluster 2 consisted of IgG targeting HCMV envelope glycoproteins, and cluster 3 comprised IgG responses against HCMV tegument proteins. Of the 9 antigen specificities tested, only total anti-UL16 IgG binding levels were significantly correlated with ADCC (rho = 0.42 p < 0.0001; **Figure 2**), and anti-UL16 IgG binding to FcγRIII V158/F158 correlated most strongly with ADCC and FcγRIII activation (**Figure 2**). Anti-UL141 IgG binding to FcγRIII V158/F158 were more modestly correlated with ADCC and FcγRIII activation (**Figure 2**). Despite clustering separately, some anti-gB, anti-gH/gL, anti-pp28, and anti-pp150 IgG FcγRIII binding responses were also modestly correlated with ADCC and/or FcγRIII activation (**Figure 2**). These weaker correlations suggest that IgG against some HCMV envelope glycoproteins or tegument proteins may help mediate ADCC, but likely to a lesser degree than UL16- and UL141-specific IgG.

**Figure 2.**
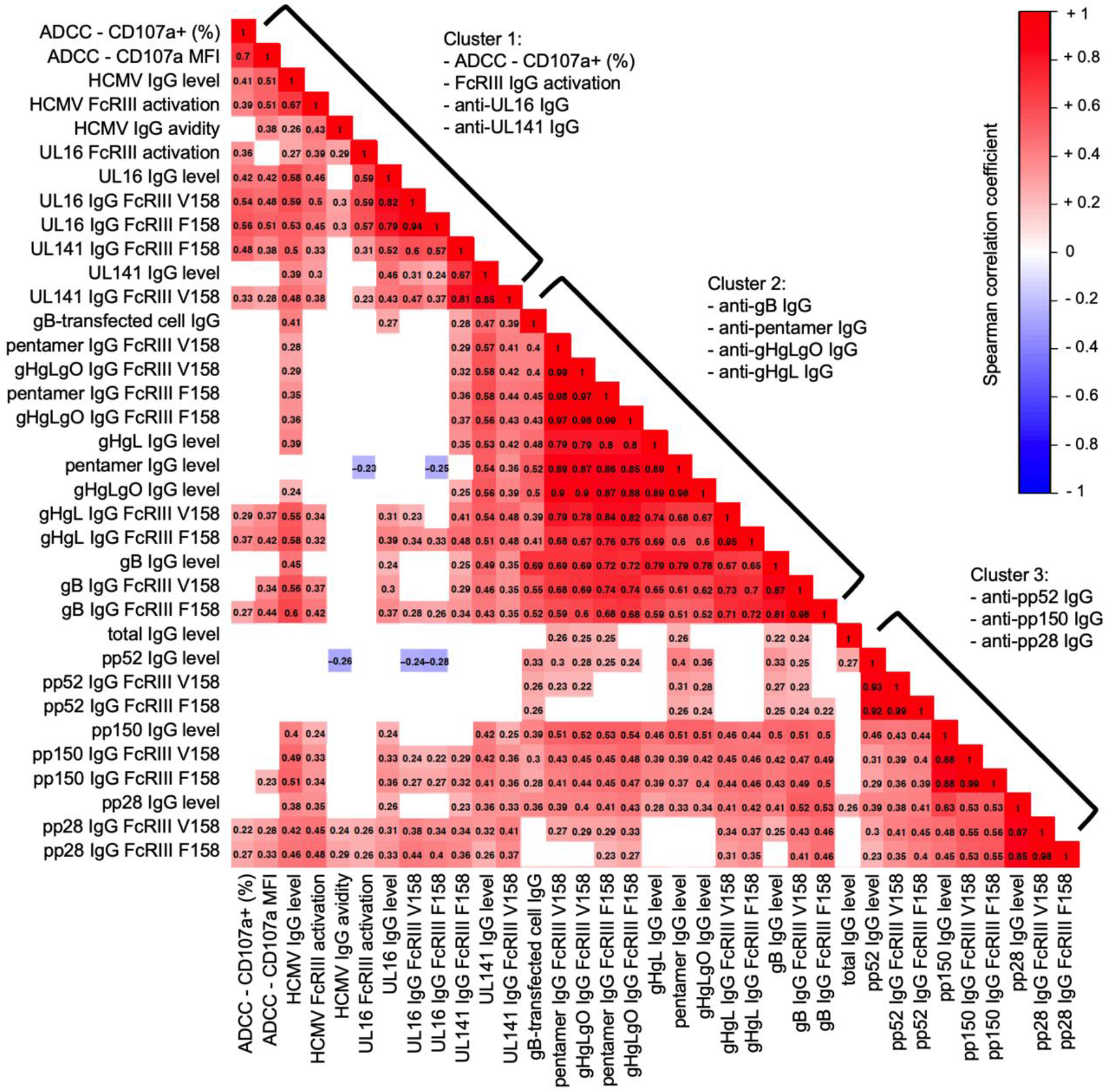
HCMV-specific NK cell ADCC and FcγRIII/CD16 activating antibodies cluster with anti-UL16 and anti-UL141 IgG responses. Hierarchical clustering was performed on Spearman correlation coefficients to group strongly correlated immune variables. Matrix of maternal sera antibody responses showing Spearman correlation coefficients from -1.0 (blue) to +1.0 (red). Non-significant correlations (*P* > 0.05) shown in white. MFI = mean florescent intensity. Level = total antigen-specific IgG binding measured by a binding antibody multiplex assay. gB-transfected cell IgG = IgG binding to cell-associated gB as measured in Semmes et al. 2022.

### Magnitude and quality of anti-HCMV IgG binding to FcγRIII differs in non-transmitting and transmitting dyads

We previously reported that anti-HCMV IgG engagement of FcγRI/FcγRIIa was enhanced in non-transmitting dyads despite having lower magnitude anti-HCMV IgG levels (7). In this follow-up study, we observed a similar phenomenon with FcγRIII engagement. Magnitude of FcγRIII binding across IgG specificities against envelope or tegument proteins was higher in transmitting versus non-transmitting dyads (**Figure 3A-B**); however, magnitude of antigen-specific IgG binding to FcγRIII incorporates both the strength of Fab binding to the antigen and the Fc-FcγRIII interaction. Since antigen-specific IgG levels differed between groups (**Supplementary Tables 1-2**), we normalized FcγRIII binding to total antigen-specific IgG levels to directly compare the quality of the Fc-FcγRIII interaction. After normalization, we found that the Fc region of anti-tegument IgG from non-transmitting dyads had better binding to both high- and low-affinity FcγRIII (**Figure 4A-B**).

**Figure 3.**
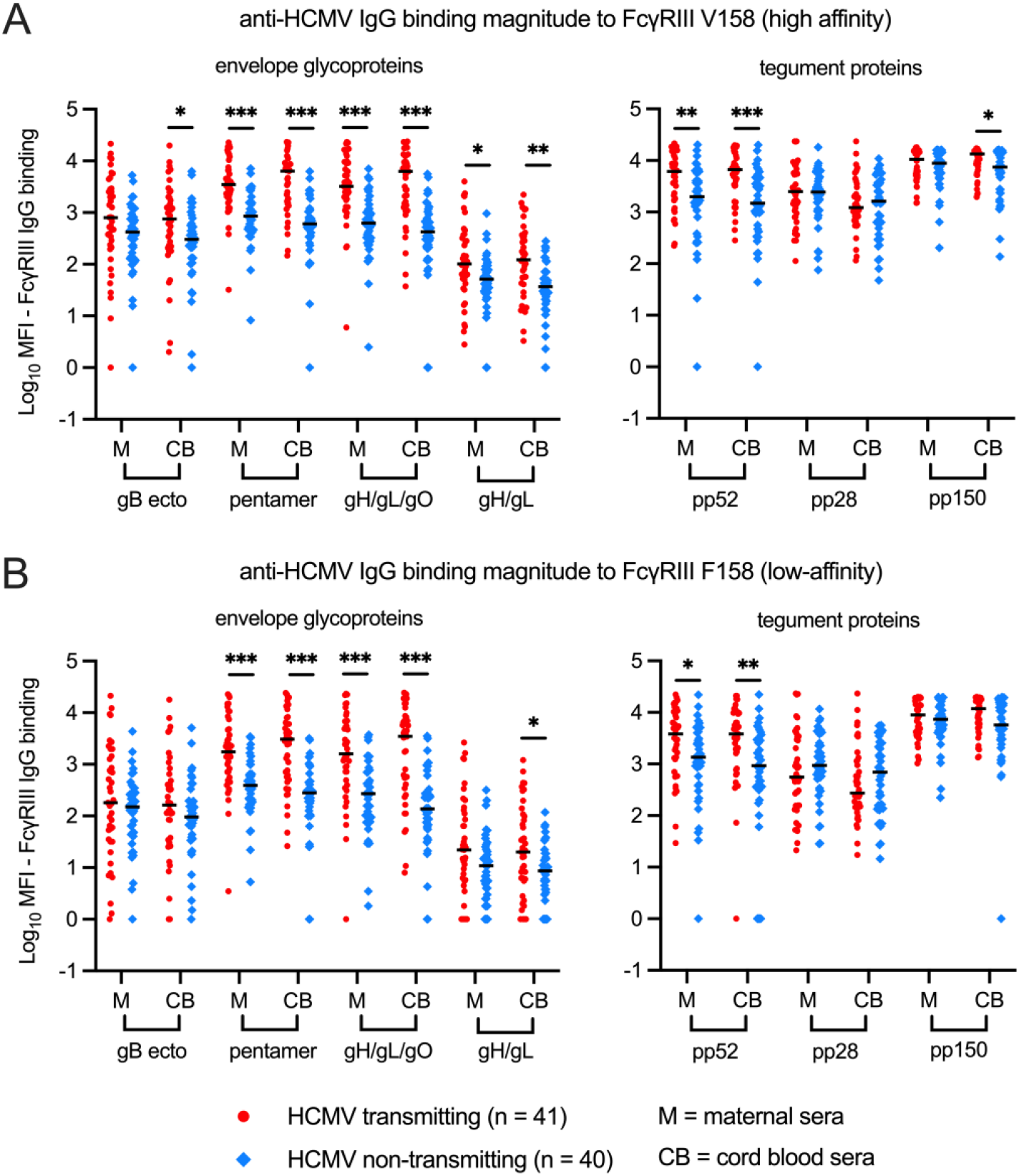
HCMV antigen-specific IgG binding magnitude to FcγRIII/CD16 in transmitting versus non-transmitting dyads. HCMV antigen-specific IgG binding to FcγRIII in maternal (M) and cord blood (CB) sera was measured using a binding antibody multiplex assay with a biotinylated FcγR and streptavidin-PE detection antibody. HCMV antigen-specific IgG binding to FcγRIII was compared between transmitting (red circles, n = 41) and non-transmitting (blue diamonds, n = 40) mother-infant dyads. (A) HCMV antigen-specific IgG binding to FcγRIII high-affinity V158 variant. (B) HCMV antigen-specific IgG binding to FcγRIII low-affinity F158 variant. Horizontal black bars denote median. FDR-corrected *P* values reported for Mann-Whitney U test. * *P* < 0.05, \*\**P* < 0.01, \*\*\**P* < 0.001.

**Figure 4.**
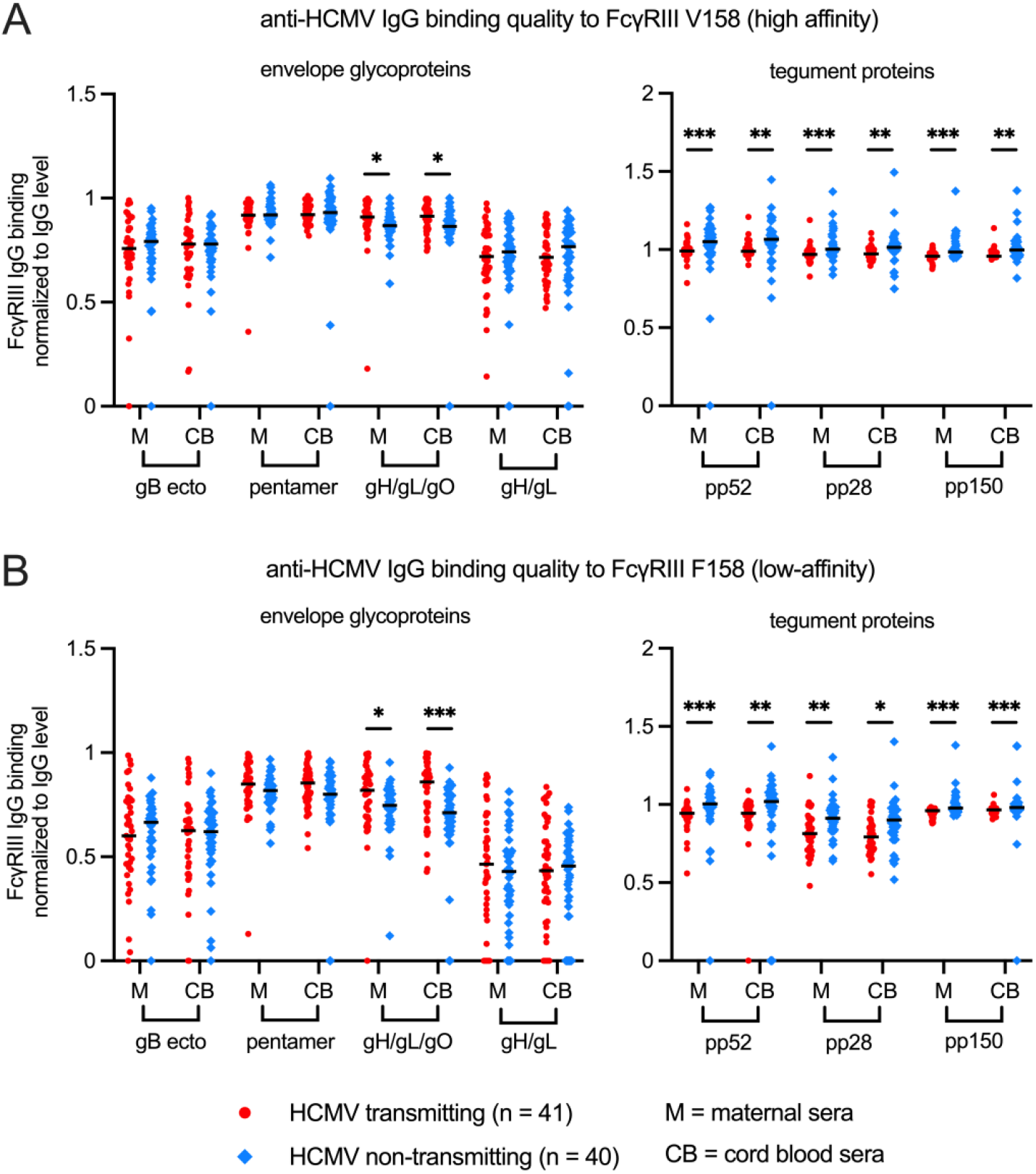
HCMV antigen-specific IgG binding quality to FcγRIII/CD16 in transmitting versus non-transmitting dyads. HCMV antigen-specific IgG binding to FcγRIII in maternal (M) and cord blood (CB) sera was measured using a binding antibody multiplex assay with a biotinylated FcγR and streptavidin-PE detection antibody. Antigen-specific IgG binding to FcγRIII was normalized to total antigen-specific IgG binding (i.e., total antigen-specific IgG level) as a ratio and compared between transmitting (red circles, n = 41) and non-transmitting (blue diamonds, n = 40) mother-infant dyads. (A) Normalized HCMV antigen-specific IgG binding to FcγRIII high-affinity V158 variant. (B) Normalized HCMV antigen-specific IgG binding to FcγRIII low-affinity F158 variant. Horizontal black bars denote median. FDR-corrected *P* values reported for Mann-Whitney U test. * *P* < 0.05, \*\**P* < 0.01, \*\*\**P* < 0.001.

### Anti-UL16, but not anti-UL141, IgG responses are associated with reduced risk of cCMV infection

Finally, we compared anti-UL141 and anti-UL16 IgG in our transmitting and non-transmitting dyads. Although modestly correlated with ADCC, anti-UL141 IgG levels were slightly higher in transmitting versus non-transmitting pregnancies (**Figure 5A, Supplementary Tables 1-2**), even though UL141-specific IgG transfer was lower in transmitting dyads (**Supplementary Figure 3A**). While the magnitude of anti-UL141 IgG binding to FcγRIII did not differ between groups, normalized anti-UL141 IgG binding to FcγRIII V158/F158 was significantly higher in non-transmitters (**Figure 5B**), suggesting better quality Fc-FcγRIII engagement. Nevertheless, anti-UL141 IgG binding was not associated with protection against transmission in our univariate regression analysis (**Tables 2-3**). In contrast, anti-UL16 IgG levels were 3-fold higher in maternal (99.8 vs. 32.3 MFI, p = 0.019) and 6-fold higher in cord blood (125.4 vs. 19.0 MFI, p = 0.002) sera from non-transmitting compared to transmitting pregnancies (**Figure 5C, Supplementary Tables 1-2**). Anti-UL16 IgG levels were lower in cord blood versus maternal sera within transmitting dyads, whereas anti-UL16 IgG levels were higher in cord blood versus maternal sera within non-transmitting dyads (**Supplementary Figure 3B**), indicating increased UL16-specific IgG transfer to uninfected infants. Anti-UL16 IgG in maternal sera of non-transmitting dyads also had 30 to 100-fold higher magnitude binding to FcγRIII V158 (420.6 vs. 13.5 MFI, p < 0.0001) and F158 (119.4 vs. 1.0 MFI, p < 0.0001) and better quality Fc-FcγRIII binding after normalization for anti-UL16 IgG levels (**Figure 5D**). Anti-UL16 IgG in maternal sera from non-transmitters also had increased functional activation of FcγRIII (p = 0.002; **Figure 5E**) when measured with our reporter cells. In our univariate regression analysis, anti-UL16 IgG binding to and activation of FcγRIII were significantly associated with reduced risk of cCMV transmission (**Tables 2-3**). When comparing anti-UL16 IgG responses in dyads stratified by RAI score and HCMV-specific IgM status, anti-UL16 IgG binding was significantly lower in dyads with low/intermediate RAI scores or detectable HCMV-specific IgM (**Supplementary Tables 3-4**). Nevertheless, anti-UL16 IgG binding to and activation of FcγRIII remained associated with protection in a sensitivity analysis excluding these dyads (**Supplementary Tables 5-6**).

**Figure 5.**
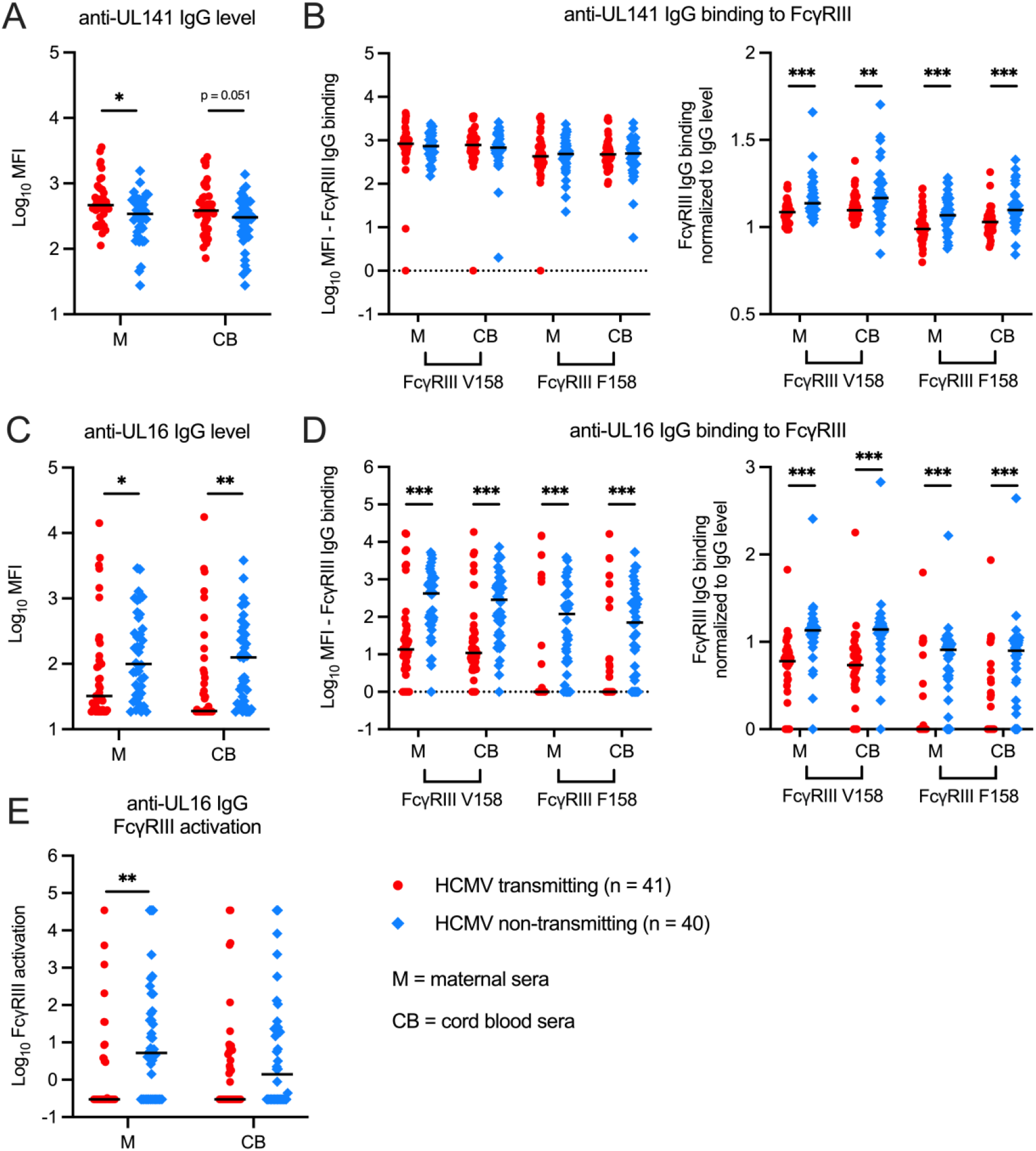
Anti-UL141 and anti-UL16 IgG binding in HCMV transmitting versus non-transmitting mother-infant dyads. Anti-UL141 and anti-UL16 IgG binding was measured with a binding antibody multiplex assay using maternal (M) and cord blood (CB) sera from HCMV transmitting (red circles, n = 41) and non-transmitting (blue diamonds, n = 40) mother-infant dyads. (A) Total anti-UL141 IgG binding (i.e., level). (B) Anti-UL141 IgG binding to FcγRIII high-affinity V158 and low-affinity F158 variants before and after normalization for total anti-UL141 IgG level. (C) Total anti-UL16 IgG binding (i.e., level). (D) Anti-UL16 IgG binding to FcγRIII high-affinity V158 and low-affinity F158 variants before and after normalization for total anti-UL16 IgG level. (E) Anti-UL16 IgG FcγRIII activation measured via FcγR-CD3ζ BW cell activation assay. FDR-corrected *P* values reported for Mann-Whitney U test. * *P* < 0.05, \*\**P* < 0.01, \*\*\**P* < 0.001.

Finally, we explored if anti-UL16 IgG may contribute to greater ADCC activation using statistical modeling. Our interaction analysis demonstrated that the association between maternal sera ADCC activation and reduced risk of cCMV infection was stronger in dyads with higher UL16-specific IgG responses (**Table 2**), indicating that anti-UL16 antibodies enhance ADCC activation in non-transmitting pregnancies. These results are underscored by visualizing the correlations between anti-UL16 IgG and ADCC activation stratified by cCMV status (**Figure 6A-B**), which highlight the strong positive relationship between these variables (p < 0.001). Taken together, our data supports a model wherein anti-UL16 IgG binding to FcγRIII on NK cells may mediate protective ADCC responses against HCMV transmission in utero (**Figure 6C**), a hypothetical mechanism of protection that should be tested in future prospective clinical cohorts and experimental studies.

**Figure 6.**
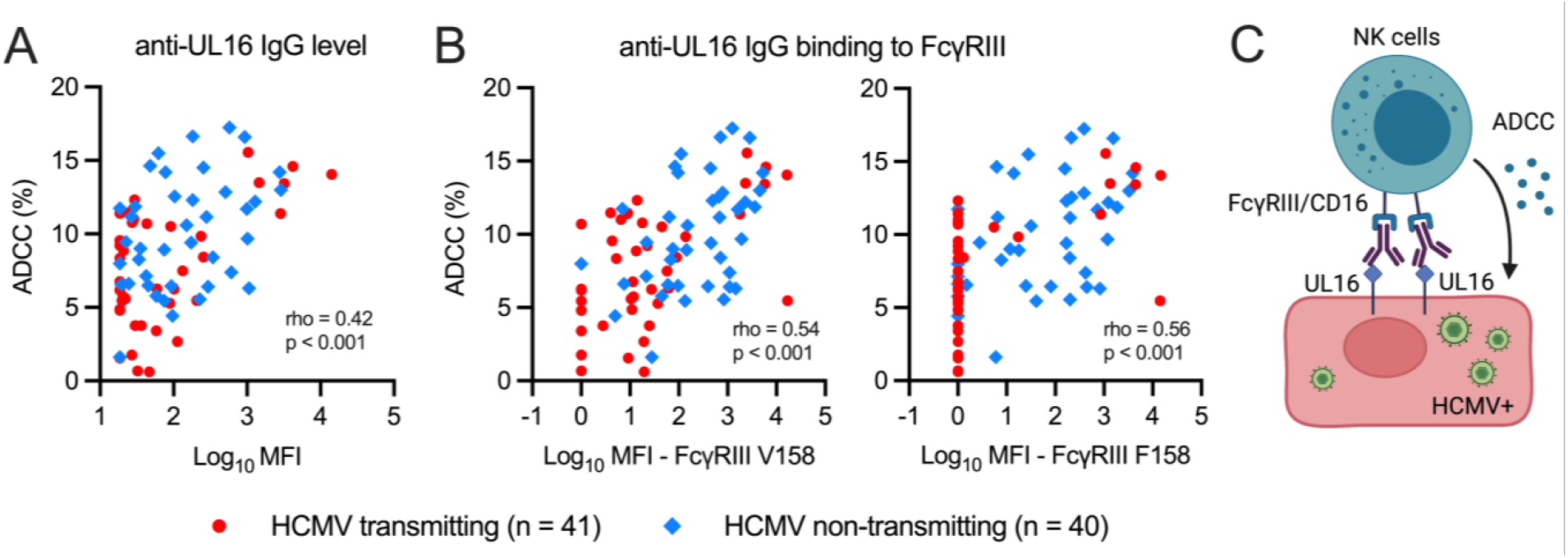
Anti-UL16 IgG binding and ADCC-activating antibodies are correlated and boosted in HCMV non-transmitting dyads. HCMV-specific ADCC (i.e., NK cell degranulation) and anti-UL16 IgG binding were measured using maternal sera from HCMV transmitting (red circles, n = 41) and non-transmitting (blue diamonds, n = 40) mother-infant dyads. (A) Scatterplots showing Spearman correlation between anti-UL16 IgG binding and ADCC. (B) Scatterplots showing Spearman correlation between ADCC and anti-UL16 IgG binding to FcγRIII high-affinity V158 and low-affinity F158 variants. (C) Hypothetical model demonstrating proposed role for anti-UL16 IgG binding and FcγRIII in mediating NK cell ADCC against HCMV-infected cells.

## DISCUSSION

Our finding that ADCC-activating antibodies are associated with protection against cCMV infection contributes to a mounting body of evidence that Fc effector functions are important for host defense against HCMV. During co-evolution with the human immune system, HCMV has developed numerous strategies to evade Fc-mediated immunity including ADCC (18). HCMV encodes multiple viral FcγR decoys that bind to host IgG to prevent FcγR engagement (19, 20, 23) and employs at least 12 viral proteins to subvert NK cell killing by engaging inhibitory receptors, removing ligands for activating receptors, and interfering with immunological synapse formation (17, 18, 21, 27, 28). Though Vlahava et al. recently demonstrated that monoclonal antibodies and pooled polyclonal sera targeting the NK cell immunoevasins UL141 and UL16 can activate ADCC (24), our study is the first to demonstrate that IgG binding against these targets correlates with FcγRIII activation and NK cell degranulation in a large mother-infant clinical cohort (n = 162 sera samples). Interestingly, anti-UL16, but not anti-UL141 IgG responses, were associated with reduced risk of cCMV infection in our cohort. Moreover, both the quantity and quality, assessed by FcγRIII engagement, of anti-UL16 IgG was greater in non-transmitting dyads. UL16 blocks ligand interactions with the host NK cell activating receptor NKG2D and is highly conserved across clinical HCMV strains (29-31). Thus, targeting UL16 to overcome NK cell immune evasion strategies and activate maternal ADCC may be a promising strategy to prevent HCMV transmission. That ADCC and anti-UL16 IgG responses were lower in dyads with low/intermediate avidity scores suggests that these responses may take significant time to develop after infection and could be primed by HCMV vaccination prior to pregnancy.

Our study has broad implications for HCMV vaccinology and immunotherapeutic development beyond congenital infection. Neutralizing antibodies against envelope glycoproteins have been a main focus of HCMV vaccines and antibody-based therapeutics (4, 32), yet there is an increasing appreciation that non-neutralizing functions i.e., antibody-mediated activation of cellular immunity are a key host defense strategy against HCMV. Several studies have demonstrated that protection from the gB/MF59 subunit vaccine, which achieved 50% efficacy in clinical trials, was mediated by non-neutralizing antibody functions, likely against the cell surface conformation of gB (15, 16, 33, 34). Antibodies elicited by the gB/MF59 vaccine stimulated robust monocyte phagocytosis but not NK cell degranulation (15, 16), suggesting that these vaccine-induced antibodies activate ADCP but not ADCC. Anti-gB antibodies likely mediate polyfunctional responses including both phagocytosis and neutralization, a phenomenon that has been observed in HIV infection (35-37); however, anti-gB IgG responses were not associated with protection in our cohort. Instead, antibodies against tegument or non-structural proteins were more clearly associated with reduced transmission risk and seemed poised to better activate FcγRs (24, 38). In particular, our study suggests that immunoevasins (e.g., UL16) should be targeted to engage both low- and high-affinity FcγRs. Taken together, our findings that antibodies stimulating ADCC and ADCP correlate with protection against cCMV infection reinforces that Fc antibody effector functions should be explored as immunologic targets against HCMV (7).

Further studies are needed to understand why certain antibodies against HCMV may engage FcγRs and activate downstream Fc effector functions better than others. Fc-mediated antibody responses are influenced by IgG Fc region characteristics such as IgG subclass and glycosylation that modify FcγR binding affinity (39-44). Our finding that the Fc region of HCMV-specific IgG in non-transmitting dyads had better quality FcγRIII binding and activation suggests that there may be differences in the Fc profiles of transmitting versus non-transmitting pregnancies. Fc region modifications to improve anti-UL16 and anti-UL141 IgG binding to FcγRIII can enhance ADCC and NK cell killing of HCMV-infected cells in vitro (24). Thus, Fc-engineering could be employed in the future to improve upon antibody-based therapeutics against HCMV (39). Modulating IgG Fc region characteristics to augment Fc engagement could improve passive immunization strategies, which are urgently needed given the lack of efficacy of HCMV hyperimmunoglobulin (HCMV-HIG) in randomized clinical trials (8, 9). It is interesting to speculate whether ineffective engagement of Fc-mediated immunity may have partially contributed to the failure of HCMV-HIG to prevent fetal transmission in prior clinical trials. We previously observed that FcγRI-mediated ADCP of HCMV was greatly reduced in the setting of high HCMV-HIG concentrations (7), leading us to hypothesize that certain Fc effector functions may have been poorly elicited in pregnant people treated with HCMV-HIG, hindering its efficacy. Overall, our work highlights that Fc characteristics should be considered when designing next generation polyclonal or monoclonal antibodies against HCMV. Novel vaccine strategies to elicit specific IgG subclasses or glycosylation profiles endogenously to enhance Fc-mediated immunity should also be explored.

The development of antibody-based prophylaxis and/or vaccines to prevent cCMV infection has been hindered by our incomplete understanding of protective maternal immunity. Moreover, whether antibodies mediate protection solely by limiting systemic maternal viral replication and placental infection or also play a role in the fetal circulation remains unclear. We found that HCMV-specific ADCC-activating antibodies were poorly transferred from maternal to cord blood sera in our cohort regardless of transmission status. In contrast, ADCC-mediating antibodies against influenza and pertussis have been shown to be robustly transferred across the placenta in healthy pregnancies (43). Since Fc region characteristics also govern transplacental IgG transport, Fc profiles likely underlie these differences in placental IgG transfer (41, 43, 44). Nevertheless, cord blood sera ADCC responses were strongly associated with protection against transmission and anti-UL16 IgG was highly transferred to uninfected infants. Vaaben et al. also recently observed that cord blood NK cells expressing FcγRIII/CD16 are expanded in utero following cCMV infection (45, 46). Taken together, these studies suggest that maternal ADCC-activating antibodies transferred across the placenta and fetal NK cells expressing FcγRIII may synergize to defend against HCMV (45, 46). Therefore, strategies to enhance placental transfer of these potentially protective antibodies through Fc-engineering and to engage fetal and/or neonatal innate immune cells in Fc-mediated immunity should be explored.

Our study is limited by its retrospective design and relatively small sample size that reduced statistical power. Due to the cross-sectional nature of this cord blood bank donor cohort, we could not identify the timing of maternal HCMV acquisition or transmission during pregnancy; thus, caution is warranted in interpreting our results since differences may be biased by a higher rate of primary infection, reinfection, and/or reactivation in transmitting versus non-transmitting dyads. This limitation and our sensitivity analyses excluding dyads with low/intermediate avidity scores or HCMV-specific IgM highlights the need for future longitudinal prospective studies to investigate protective immunity against cCMV in maternal primary and non-primary infection. Since all infants in the study were born healthy, we could not assess clinical correlations between antibody responses and symptomatic versus asymptomatic cCMV infection, which may introduce some skewing in our results. Longitudinal clinical data such as neurodevelopmental outcomes and hearing loss were not collected retrospectively, so it is unknown if any of the cCMV cases had delayed sequalae. Sera sample volumes were very limited, preventing additional mechanistic experiments and more work is needed to define the anti-viral functions of ADCC/FcγRIII activation and anti-UL16 antibodies in controlling viral replication. Maternal PBMCs were not collected, so we were also unable to assess cellular correlates of protection. Whether maternal NK cell abundance, phenotype, or function differs in transmitting and non-transmitting pregnancies should be investigated.

In conclusion, our study indicates that ADCC and FcγRIII activating antibodies against HCMV and IgG against the NK cell immunoevasion protein UL16 may help protect against cCMV infection. Our work suggests that designing HCMV vaccines or antibody-based therapeutics that can engage FcγRs and overcome NK cell immune evasion strategies may be an effective approach to combating this ubiquitous herpesvirus that is the leading infectious cause of congenital disease and disability worldwide.

## METHODS

### Study population

We analyzed maternal (n = 81) and cord blood (n = 81) sera samples from a retrospective cohort of mother-infant donors to the Carolinas Cord Blood Bank (CCBB), which has been previously described in our recent complementary study (**Supplementary Figure 1**) (7). All mothers in our study were HCMV IgG seropositive and cases of cCMV infection were identified by HCMV DNAemia, detected by PCR, in the cord blood plasma at birth. “HCMV transmitting” cases with cCMV infection (n=41) were matched to a target of 1 “HCMV non-transmitting” mother-infant dyad (n=40). Maternal HCMV IgG seropositivity was confirmed by a whole-virion HCMV ELISA and HCMV IgM seropositivity was determined using a clinical diagnostic ELISA (Bio-Rad CMV IgM EIA Kit). HCMV-specific IgG relative avidity index (RAI) scores were determined by calculating the mean RAI across 3 HCMV trains (TB40/E, AD169r, and Toledo virus) using whole virion ELISA using urea as the dissociation agent as previously described (7). Maternal RAI scores < 60% were defined as low/intermediate avidity and >= to 60% were defined as high avidity (25). Matching criteria included infant sex, infant race, maternal age, and delivery year. Only people with healthy, uncomplicated pregnancies that gave birth at term were included in our study; cord blood donors were screened for signs of (a) neonatal sepsis, (b) congenital infection (petechial rash, thrombocytopenia, hepatosplenomegaly), and (c) congenital abnormalities.

### Natural killer (NK) cell-mediated ADCC

NK cell degranulation was quantified by cell-surface expression of CD107a as previously described (47). MRC-5 fibroblasts (target cells; ATCC) were infected with HCMV strain AD169r (a AD169 derivative with repaired UL128-131 expression named BadrUL131-Y4-GFP (48)) at an MOI of 1.0 or mock-infected. After 48 hours, primary human NK cells (effector cells) were isolated by negative selection with magnetic beads (human NK cell isolation kit, Miltenyi Biotech) from PBMCs of a healthy adult donor then live, primary NK cells were added to each well containing HCMV-infected or mock-infected fibroblasts at an E:T ratio of 1:1. Cytogam IgG product or diluted sera samples (1:75) were then added with brefeldin A (GolgiPlug; BD Biosciences), monensin (GolgiStop; BD Biosciences), and anti-CD107a–FITC (clone H4A3; BD Biosciences). After a 6-hour incubation, NK cells were stained with anti-CD56-PE/Cy7 (clone NCAM16.2; BD Biosciences), anti-CD16-PacBlue (clone 3G8; BD Biosciences), and a viability dye (Live/Dead Aqua Dead Cell Stain, ThermoFisher). Events were acquired on an LSR Fortessa flow cytometer, and the frequencies of live, CD107a positive NK cells were calculated in FlowJo (gating strategy in **Supplementary Figure 2**). To correct for non-specific degranulation activity, the signal in mock-infected wells was subtracted from the signal in HCMV-infected wells for each sera sample (**Supplementary Figure 2**).

### HCMV antigen-specific IgG binding

IgG binding to HCMV antigens including UL16 and UL141 (provided by R. Stanton), envelope glycoproteins (gB ectodomain, pentamer complex, gH/gL/gO, gH/gL), and tegument proteins (pp28, pp150, pp52) was quantified using a binding antibody multiplex assay (BAMA) as previously described (7). In brief, HCMV antigens were coupled to intrinsically fluorescent beads (Bio-Plex pro magnetic COOH beads, Bio-Rad) then co-incubated with serially diluted Cytogam IgG product (CSL Behring) or sera samples. Antigen-specific IgG binding was detected with mouse anti-human IgG-PE (Southern Biotech) and MFI was acquired on a Bio-Plex 200.

### HCMV antigen-specific IgG binding to FcγRs

Antigen-specific IgG binding to FcγRIII/CD16 was measured using a modified BAMA as previously described (7). Purified human FcγRIII high-affinity (V158) and low-affinity (F158) variants were produced by the DHVI Protein Production Facility and biotinylated in-house. First, sera samples were co-incubated with HCMV antigen-coated beads, as above. Next, biotinylated FcγRIII was complexed with streptavidin-PE (BD Biosciences) then co-incubated with antibody-bound beads and MFI was acquired on a Bio-Plex 200.

### FcγR IgG activation

HCMV-specific IgG activation of FcγRIII/CD16 was quantified using mouse BW thymoma cells expressing chimeric FcγR-CD3ζ as previously described (7, 49). To confirm FcγR expression, BW cells were stained with anti-FcγRI/CD64-PE (clone 10.1, eBioscience), anti-FcγRII/CD32-PE (clone 6C4, eBioscience), anti-FcγRIII/CD16-PE (clone CB16, eBioscience), and anti-Ig-PE isotype control (eBioscience). Events were acquired on a LSRII flow cytometer then analyzed using FlowJo. To quantify FcγR activation, 96-well plates were coated with HCMV strain AD169r (20,000 PFU/well) or UL16 antigen (250 μg/well) then co-incubated with Cytogam IgG product or sera samples (diluted 1:10) to form immune complexes. Next, FcγRIII-expressing BW cells were added and incubated for 20 hours. Cell supernatants were then harvested, and mouse IL-2 levels were quantified as a read-out of FcγRIII-CD3ζ activation using ELISA as previously described (7).

### Statistics

All primary data underwent independent quality control by another lab member using standardized criteria base on duplicate well variance and performance of positive and negative controls, which included Cytogam, HCMV seropositive, and HCMV seronegative sera samples. Wilcoxon rank-sum tests were used to compare transmitting and non-transmitting dyads and Wilcoxon signed rank tests were used to assess differences within dyads. Spearman’s correlation coefficient was calculated for select immune variables and correlation matrices were plotted using the corrplot package in R v4.1.

ELISA and BAMA data were log-transformed for all regression analyses. Statistical significance was defined a priori as p < 0.05 after an FDR correction for multiple comparisons. Statistical analyses were completed in R v4.1 and GraphPad Prism v9.1.

### Study Approval

Approval was obtained from Duke University School of Medicine’s Institutional Review Board (Pro00089256) to use deidentified clinical data and biospecimens provided by the CCBB. No patients were prospectively recruited for this study, and all samples were acquired retrospectively from the CCBB biorepository from donors who had previously provided written consent for banked biospecimens to be used for research.

## Data Availability

All data produced in the present study are available upon reasonable request to the authors.

## Author contributions

ECS, JP, and SRP designed the research study. ECS, IGM, NR, and CP conducted the experiments and acquired the data. ECS, IGM, and NR completed the primary data analysis. ECS completed the statistical analyses with oversight from KMW. SRP and KMW acquired funding for the study. JHH helped acquire the human samples and RS provided key reagents for the study. ECS wrote the primary draft of the manuscript. ECS, IGM, NR, CP, JHH, KMW, JP, RS and SRP all contributed to writing and editing the manuscript.

## Acknowledgements

Thank you to the CCBB donors, CCBB director Dr. Joanne Kurtzberg, and CCBB staff including Jose Hernandez, Ann Kaestner, and Korrynn Vincent who were instrumental in acquiring the biospecimens and donor clinical information for this study. We also would like to thank Drs. Phillip Kolb and Hartmut Hengel who generously provided the FcR-transfected BW cell lines. This project was supported by NIH NCI 1R21CA242439-01 “Immune Correlates and Mechanisms of Perinatal Cytomegalovirus Infection and Later Life ALL Development” to KMW and SRP and NIH NIAID 1R21-AI147992 “Humoral immune correlates of protection against congenital CMV and HSV transmission in HIV-infected women” to SRP. This project was also partially sponsored by Moderna. The funders had no role in study design, data collection and analysis, decision to publish, or preparation of the manuscript.

