## Supplementary Figures and Tables for "ADCC-activating antibodies correlate with protection against congenital human cytomegalovirus infection"

**Supplementary Figure 1. Identification of HCMV transmitting and non-transmitting mother-infant dyads from the Carolinas Cord Blood Bank (CCBB) biorepository.**

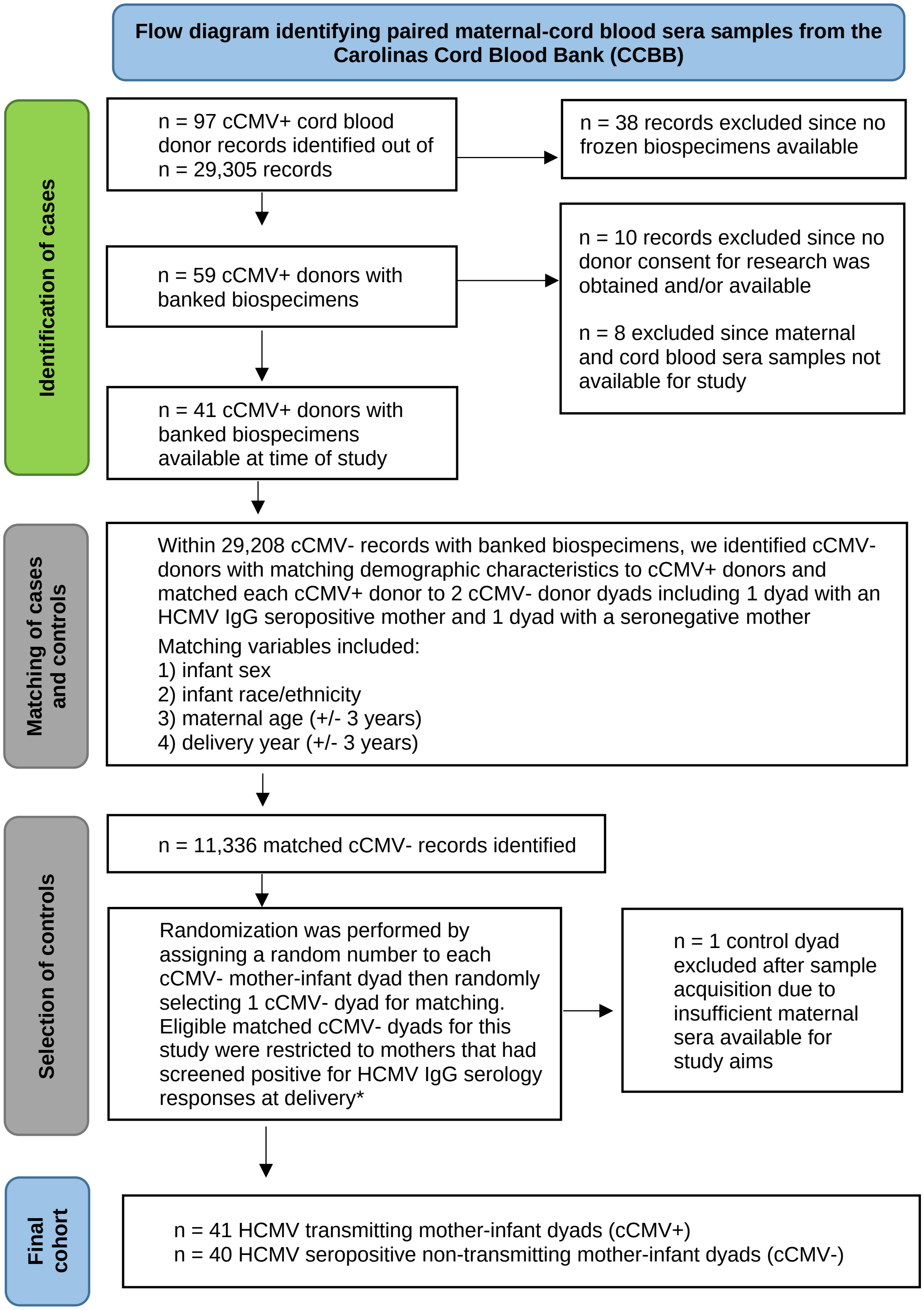


cCMV = congenital HCMV infection
cCMV+ = positive HCMV PCR cord blood screening at birth
cCMV- = negative HCMV PCR cord blood screening at birth

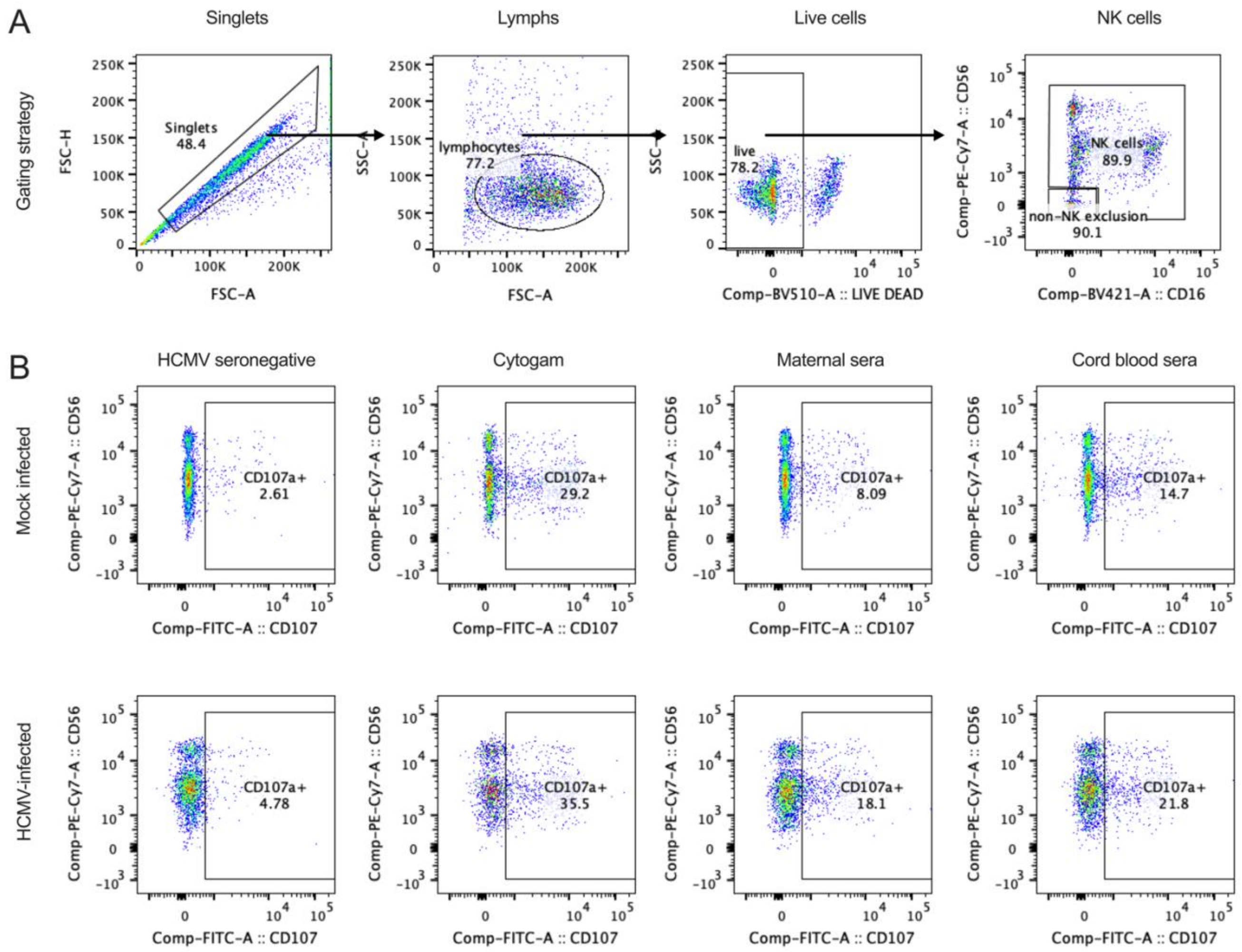


**Supplementary Figure 2. NK cell degranulation gating strategy for ADCC assay.** Antibody-dependent cellular cytotoxicity (ADCC) was measured by quantifying NK cell degranulation (i.e., CD107a positivity) against HCMV-infected fibroblasts. PBMCs were rested overnight then primary NK cells were isolated by negative selection with magnetic beads prior to co-incubation with HCMV-infected and mock-infected cells. (A) Gating strategy: singlets → lymphocytes → live lymphocytes → NK cells → CD107a positive cells. (B) CD107a positive cells were identified in the NK cell gate under mock-infected and HCMV-infected (MOI = 1) conditions for HCMV seronegative, Cytogam, and HCMV seropositive maternal and cord blood sera samples. HCMV-specific ADCC activating antibody responses were calculated by subtracting the % CD107a positive cells in the mock-infected condition from the % CD107a positive cells in the HCMV-infected condition for each sample.

| **Supplementary Table 1. Maternal sera HCMV-specific antibody responses in HCMV transmitting versus non-transmitting dyads** | | | | | | |
|---|---|---|---|---|---|---|
| | **HCMV transmitting (n = 41)** | | **HCMV non-transmitting (n = 40)** | | | |
| **Variable** | **Median** | **IQR** | **Median** | **IQR** | **p value** | **FDR p value** |
| HCMV ADCC | 7.5 | 5.7 | 9.6 | 6.0 | **0.012** | **0.020** |
| HCMV IgG FcγRIII activation | 9481.4 | 7494.3 | 12864.4 | 13557.8 | 0.054 | 0.076 |
| UL16 IgG binding level | 32.3 | 92.6 | 99.8 | 342.8 | **0.019** | **0.029** |
| UL16 IgG FcγRIII V158 | 13.5 | 53.5 | 420.6 | 1028.1 | **<0.001** | **<0.001** |
| UL16 IgG FcγRIII F158 | 1.0 | 0.0 | 119.4 | 391.2 | **<0.001** | **<0.001** |
| UL16 IgG FcγRIII activation | 0.3 | 2.8 | 5.2 | 61.3 | **0.002** | **0.005** |
| UL141 IgG binding level | 465.0 | 460.0 | 341.9 | 351.7 | **0.010** | **0.017** |
| UL141 IgG FcγRIII V158 | 833.0 | 480.3 | 736.7 | 770.4 | 0.360 | 0.440 |
| UL141 IgG FcγRIII F158 | 431.5 | 439.5 | 484.9 | 411.1 | 0.640 | 0.690 |
| gB-transfected cell IgG binding | 24.4 | 2.7 | 21.1 | 7.1 | **0.005** | **0.009** |
| gB IgG binding level | 6917.5 | 9747.1 | 1591.3 | 3126.6 | **<0.001** | **0.001** |
| gB IgG FcγRIII V158 | 798.5 | 2605.7 | 418.3 | 709.8 | **0.049** | 0.070 |
| gB IgG FcγRIII F158 | 181.5 | 1237.2 | 150.4 | 253.8 | 0.290 | 0.370 |
| pentamer IgG binding level | 7788.8 | 12098.0 | 1471.8 | 2600.2 | **<0.001** | **<0.001** |
| pentamer FcγRIII V158 | 3506.5 | 5463.3 | 854.6 | 1138.3 | **<0.001** | **<0.001** |
| pentamer IgG FcγRIII F158 | 1746.0 | 4114.3 | 391.9 | 450.5 | **<0.001** | **<0.001** |
| gHgLgO IgG binding level | 9468.0 | 14464.4 | 1666.9 | 3341.4 | **<0.001** | **<0.001** |
| gHgLgO FcγRIII V158 | 3235.3 | 7297.0 | 630.3 | 976.7 | **<0.001** | **<0.001** |
| gHgLgO IgG FcγRIII F158 | 1598.0 | 4973.8 | 270.7 | 612.2 | **<0.001** | **<0.001** |
| gHgL IgG binding level | 704.8 | 893.2 | 213.2 | 296.2 | **<0.001** | **<0.001** |
| gHgL FcγRIII V158 | 102.0 | 270.5 | 51.4 | 64.2 | **0.009** | **0.016** |
| gHgL IgG FcγRIII F158 | 22.0 | 106.0 | 10.9 | 19.5 | 0.094 | 0.130 |
| pp52 IgG binding level | 6774.3 | 9980.0 | 1077.5 | 2805.6 | **<0.001** | **<0.001** |
| pp52 FcγRIII V158 | 6120.0 | 10500.3 | 1982.3 | 4796.3 | **0.003** | **0.006** |
| pp52 IgG FcγRIII F158 | 3839.3 | 8998.2 | 1351.2 | 2793.8 | **0.011** | **0.018** |
| pp28 IgG binding level | 2606.8 | 5139.5 | 1689.3 | 3689.4 | 0.071 | 0.097 |
| pp28 FcγRIII V158 | 2503.5 | 4755.5 | 2454.5 | 3950.9 | 0.980 | 0.980 |
| pp28 IgG FcγRIII F158 | 557.5 | 3223.8 | 934.2 | 2410.5 | 0.390 | 0.460 |
| pp150 IgG binding level | 16009.3 | 17044.8 | 7059.9 | 14579.7 | **0.006** | **0.011** |
| pp150 FcγRIII V158 | 10488.3 | 10381.3 | 8811.8 | 8544.8 | 0.360 | 0.440 |
| pp150 IgG FcγRIII F158 | 9012.8 | 14228.7 | 7410.2 | 9211.9 | 0.500 | 0.560 |
| Bold indicates statistical significance ($p < 0.05$). | | | | | | |

| Supplementary Table 2. Cord blood sera HCMV-specific antibody responses in HCMV transmitting versus non-transmitting dyads | | | | | | |
|---|---|---|---|---|---|---|
| | HCMV transmitting (n = 41) | | HCMV non-transmitting (n = 40) | | | |
| **Variable** | **Median** | **IQR** | **Median** | **IQR** | **p value** | **FDR p value** |
| HCMV ADCC | 4.8 | 3.8 | 7.1 | 4.0 | **0.001** | **0.003** |
| HCMV IgG FcγRIII activation | 8790.8 | 7983.3 | 12847.7 | 14424.3 | **0.008** | **0.015** |
| UL16 IgG binding level | 19.0 | 54.4 | 125.4 | 278.5 | **0.002** | **0.004** |
| UL16 IgG FcγRIII V158 | 11.0 | 37.3 | 284.3 | 754.9 | **<0.001** | **<0.001** |
| UL16 IgG FcγRIII F158 | 1.0 | 4.3 | 70.8 | 265.2 | **<0.001** | **<0.001** |
| UL16 IgG FcγRIII activation | 0.3 | 6.5 | 1.4 | 21.0 | 0.400 | 0.470 |
| UL141 IgG binding level | 385.5 | 361.2 | 302.0 | 352.9 | **0.035** | 0.051 |
| UL141 IgG FcγRIII V158 | 783.5 | 644.5 | 677.3 | 771.5 | 0.420 | 0.490 |
| UL141 IgG FcγRIII F158 | 476.3 | 354.0 | 496.3 | 446.6 | 0.970 | 0.980 |
| gB-transfected cell IgG binding | 24.0 | 2.1 | 21.8 | 6.7 | **0.002** | **0.003** |
| gB IgG binding level | 4733.0 | 7638.2 | 1824.2 | 1945.2 | **<0.001** | **0.001** |
| gB IgG FcγRIII V158 | 750.8 | 1714.5 | 305.1 | 441.3 | **0.016** | **0.026** |
| gB IgG FcγRIII F158 | 163.8 | 446.7 | 95.9 | 156.6 | 0.200 | 0.260 |
| pentamer IgG binding level | 10772.1 | 16600.4 | 952.1 | 1904.4 | **<0.001** | **<0.001** |
| pentamer FcγRIII V158 | 6350.3 | 7475.2 | 606.6 | 751.5 | **<0.001** | **<0.001** |
| pentamer IgG FcγRIII F158 | 3059.0 | 5823.8 | 281.7 | 335.8 | **<0.001** | **<0.001** |
| gHgLgO IgG binding level | 10785.4 | 18236.0 | 973.3 | 2280.3 | **<0.001** | **<0.001** |
| gHgLgO FcγRIII V158 | 6267.8 | 10928.2 | 424.9 | 741.7 | **<0.001** | **<0.001** |
| gHgLgO IgG FcγRIII F158 | 3482.8 | 6573.0 | 138.0 | 218.3 | **<0.001** | **<0.001** |
| gHgL IgG binding level | 772.3 | 1562.0 | 123.6 | 225.3 | **<0.001** | **<0.001** |
| gHgL FcγRIII V158 | 122.3 | 313.2 | 37.2 | 49.5 | **0.001** | **0.002** |
| gHgL IgG FcγRIII F158 | 20.0 | 91.8 | 8.7 | 12.9 | **0.017** | **0.027** |
| pp52 IgG binding level | 6894.0 | 7742.3 | 634.3 | 2465.6 | **<0.001** | **<0.001** |
| pp52 FcγRIII V158 | 6641.0 | 7955.7 | 1486.8 | 4117.8 | **<0.001** | **<0.001** |
| pp52 IgG FcγRIII F158 | 3809.0 | 8143.0 | 922.7 | 2610.6 | **0.001** | **0.002** |
| pp28 IgG binding level | 1459.0 | 1972.8 | 1383.7 | 2528.4 | 0.310 | 0.390 |
| pp28 FcγRIII V158 | 1226.5 | 2212.0 | 1623.0 | 3973.5 | 0.880 | 0.910 |
| pp28 IgG FcγRIII F158 | 275.0 | 989.2 | 697.9 | 2428.9 | 0.370 | 0.440 |
| pp150 IgG binding level | 20199.0 | 14946.0 | 6307.0 | 15201.4 | **0.001** | **0.003** |
| pp150 FcγRIII V158 | 13487.3 | 8827.5 | 7418.3 | 11574.3 | **0.020** | **0.030** |
| pp150 IgG FcγRIII F158 | 11753.8 | 12327.8 | 5710.8 | 10773.8 | **0.029** | **0.042** |
| Bold indicates statistical significance ($p < 0.05$). | | | | | | |

| Supplementary Table 3. Maternal sera antibody responses in dyads with low/intermediate versus high HCMV IgG avidity scores [a] | | | | | | |
|---|---|---|---|---|---|---|
| | Low/intermediate RAI score (n = 9) | | High RAI score (n = 72) | | | |
| **Variable** | **Median** | **IQR** | **Median** | **IQR** | **p value** | **FDR p value** |
| HCMV ADCC | 5.1 | 2.6 | 9.5 | 5.8 | **0.001** | **0.004** |
| HCMV IgG FcγRIII activation | 7041.7 | 1067.1 | 12517.9 | 11502.6 | **<0.001** | **0.002** |
| UL16 IgG binding level | 20.2 | 13.2 | 90.9 | 259.2 | **0.002** | **0.008** |
| UL16 IgG FcγRIII V158 | 1.9 | 9.2 | 90.3 | 949.4 | **<0.001** | **0.001** |
| UL16 IgG FcγRIII F158 | 1.0 | 0.0 | 11.8 | 389.1 | **0.003** | **0.011** |
| UL16 IgG FcγRIII activation | 0.3 | 0.0 | 2.6 | 35.2 | **0.022** | **0.041** |
| UL141 IgG binding level | 296.5 | 239.3 | 419.5 | 330.1 | 0.130 | 0.170 |
| UL141 IgG FcγRIII V158 | 507.0 | 498.5 | 850.8 | 772.9 | **0.004** | **0.012** |
| UL141 IgG FcγRIII F158 | 189.3 | 186.9 | 488.3 | 471.5 | **<0.001** | **0.001** |
| gB-transfected cell IgG binding | 23.0 | 4.5 | 23.8 | 4.8 | 0.550 | 0.600 |
| gB IgG binding level | 2740.6 | 3899.8 | 3637.0 | 7335.6 | 0.470 | 0.530 |
| gB IgG FcγRIII V158 | 292.4 | 419.9 | 651.0 | 1625.3 | 0.100 | 0.140 |
| gB IgG FcγRIII F158 | 42.7 | 137.1 | 191.8 | 636.5 | **0.041** | 0.065 |
| pentamer IgG binding level | 5601.5 | 8726.1 | 3709.0 | 7396.3 | 0.550 | 0.600 |
| pentamer FcγRIII V158 | 1999.8 | 2860.0 | 1896.0 | 3028.6 | 0.840 | 0.870 |
| pentamer IgG FcγRIII F158 | 762.3 | 1864.7 | 651.8 | 1922.8 | 0.790 | 0.830 |
| gHgLgO IgG binding level | 4950.7 | 10157.5 | 4640.0 | 8556.6 | 0.960 | 0.960 |
| gHgLgO FcγRIII V158 | 1369.5 | 3219.3 | 1750.0 | 3651.3 | 0.900 | 0.920 |
| gHgLgO IgG FcγRIII F158 | 313.7 | 1349.5 | 716.0 | 2401.6 | 0.570 | 0.610 |
| gHgL IgG binding level | 249.2 | 330.3 | 447.5 | 689.6 | 0.200 | 0.250 |
| gHgL FcγRIII V158 | 33.9 | 109.5 | 68.8 | 110.6 | 0.140 | 0.180 |
| gHgL IgG FcγRIII F158 | 2.8 | 22.6 | 14.8 | 42.8 | 0.070 | 0.100 |
| pp52 IgG binding level | 10843.7 | 13307.5 | 2494.5 | 5583.1 | **0.010** | **0.022** |
| pp52 FcγRIII V158 | 6891.3 | 12057.0 | 3681.0 | 6418.1 | 0.140 | 0.180 |
| pp52 IgG FcγRIII F158 | 4038.3 | 8863.0 | 1874.8 | 4564.4 | 0.240 | 0.300 |
| pp28 IgG binding level | 1787.3 | 1468.2 | 2465.3 | 4081.5 | 0.280 | 0.340 |
| pp28 FcγRIII V158 | 1079.7 | 1475.8 | 2537.3 | 5050.5 | **0.028** | **0.049** |
| pp28 IgG FcγRIII F158 | 220.3 | 349.1 | 888.3 | 3529.0 | **0.008** | **0.019** |
| pp150 IgG binding level | 11497.6 | 17510.0 | 10391.3 | 18133.3 | 0.950 | 0.960 |
| pp150 FcγRIII V158 | 4789.9 | 8643.3 | 9824.5 | 9385.9 | 0.097 | 0.140 |
| pp150 IgG FcγRIII F158 | 4026.3 | 6731.6 | 8581.0 | 12943.8 | 0.061 | 0.092 |

[a] HCMV IgG relative avidity index (RAI) score was calculated as the mean RAI measured against 3 HCMV strains. RAI scores <60% were considered low/intermediate avidity whereas scores ≥60% were considered high avidity. Bold indicates statistical significance ($p < 0.05$).

| Supplementary Table 4. Maternal sera antibody responses in dyads with and without detectable HCMV-specific IgM [a] | | | | | | |
|---|---|---|---|---|---|---|
| | HCMV IgM+ mothers (n=13) | | HCMV IgM- mothers (n=68) | | | |
| **Variable** | **Median** | **IQR** | **Median** | **IQR** | **p value** | **FDR p value** |
| HCMV ADCC | 6.8 | 8.0 | 9.1 | 5.7 | 0.130 | 0.240 |
| HCMV IgG FcγRIII activation | 7717.6 | 5689.2 | 12136.6 | 11592.6 | **0.044** | 0.120 |
| UL16 IgG binding level | 29.5 | 27.3 | 89.9 | 256.7 | **0.026** | 0.090 |
| UL16 IgG FcγRIII V158 | 9.0 | 18.3 | 93.6 | 884.3 | **0.001** | **0.017** |
| UL16 IgG FcγRIII F158 | 1.0 | 0.0 | 12.9 | 388.4 | **0.002** | **0.022** |
| UL16 IgG FcγRIII activation | 0.3 | 0.0 | 2.0 | 31.6 | 0.076 | 0.180 |
| UL141 IgG binding level | 376.3 | 355.7 | 417.2 | 339.7 | 0.860 | 0.920 |
| UL141 IgG FcγRIII V158 | 669.3 | 506.5 | 838.2 | 780.6 | 0.084 | 0.180 |
| UL141 IgG FcγRIII F158 | 296.8 | 286.5 | 484.9 | 498.4 | **0.025** | 0.090 |
| gB-transfected cell IgG binding | 22.9 | 4.7 | 23.8 | 4.8 | 0.900 | 0.960 |
| gB IgG binding level | 4245.5 | 11222.5 | 2762.0 | 5690.9 | 0.620 | 0.750 |
| gB IgG FcγRIII V158 | 1262.0 | 1862.0 | 477.4 | 1047.3 | 0.460 | 0.610 |
| gB IgG FcγRIII F158 | 445.0 | 692.7 | 158.1 | 364.7 | 0.590 | 0.730 |
| pentamer IgG binding level | 7703.5 | 15607.6 | 3711.1 | 6498.0 | 0.084 | 0.180 |
| pentamer FcγRIII V158 | 3203.5 | 5246.5 | 1572.8 | 2766.1 | 0.073 | 0.180 |
| pentamer IgG FcγRIII F158 | 2225.5 | 4176.2 | 611.8 | 1502.8 | 0.130 | 0.240 |
| gHgLgO IgG binding level | 9859.8 | 15935.0 | 4208.0 | 7848.2 | 0.077 | 0.180 |
| gHgLgO FcγRIII V158 | 4511.0 | 5208.2 | 1235.8 | 2634.4 | 0.061 | 0.160 |
| gHgLgO IgG FcγRIII F158 | 2445.5 | 4781.0 | 433.7 | 1354.2 | 0.091 | 0.190 |
| gHgL IgG binding level | 509.5 | 580.0 | 410.9 | 664.8 | 0.750 | 0.830 |
| gHgL FcγRIII V158 | 63.8 | 266.5 | 69.2 | 98.2 | 0.550 | 0.700 |
| gHgL IgG FcγRIII F158 | 7.0 | 63.3 | 15.1 | 32.5 | 0.340 | 0.470 |
| pp52 IgG binding level | 11638.8 | 14187.0 | 2429.8 | 5181.9 | **0.003** | **0.023** |
| pp52 FcγRIII V158 | 7662.5 | 9836.8 | 3031.6 | 5465.0 | **0.014** | 0.063 |
| pp52 IgG FcγRIII F158 | 4913.3 | 8810.5 | 1491.6 | 4591.1 | **0.022** | 0.085 |
| pp28 IgG binding level | 2606.8 | 5061.5 | 2209.0 | 3962.8 | 0.290 | 0.420 |
| pp28 FcγRIII V158 | 1690.3 | 4247.3 | 2466.7 | 4595.6 | 0.990 | 0.990 |
| pp28 IgG FcγRIII F158 | 555.3 | 2684.5 | 854.9 | 3196.8 | 0.650 | 0.780 |
| pp150 IgG binding level | 23035.8 | 11150.5 | 9472.6 | 17520.5 | **0.016** | 0.069 |
| pp150 FcγRIII V158 | 14658.5 | 10975.8 | 9216.9 | 9331.6 | 0.190 | 0.310 |
| pp150 IgG FcγRIII F158 | 14402.5 | 14151.7 | 7566.6 | 9674.8 | 0.240 | 0.360 |
| Bold indicates statistical significance ($p < 0.05$). | | | | | | |

**Supplementary Table 5. Univariate logistic regression analysis of antibody responses and risk of cCMV infection excluding HCMV IgG low/intermediate avidity mothers**

| Antibody variable | Maternal sera | | | Cord blood sera | | |
|---|---|---|---|---|---|---|
| | OR[a] | 95% CI | p value | OR | 95% CI | p value |
| **HCMV ADCC** | 0.89 | 0.78 - 1.01 | 0.065 | 0.81 | 0.68 - 0.96 | **0.017** |
| **HCMV IgG FcγRIII activation** | 1.09 | 0.73 - 1.65 | 0.658 | 1.09 | 0.76 - 1.59 | 0.632 |
| **UL16 IgG binding level level** | 0.87 | 0.65 - 1.16 | 0.341 | 0.83 | 0.62 - 1.12 | 0.228 |
| **UL16 IgG FcγRIII V158** | 0.76 | 0.61 - 0.94 | **0.010** | 0.70 | 0.56 - 0.88 | **0.003** |
| **UL16 IgG FcγRIII F158** | 0.79 | 0.67 - 0.94 | **0.006** | 0.75 | 0.62 - 0.90 | **0.003** |
| **UL16 IgG FcγRIII activation** | 0.85 | 0.73 - 1.00 | **0.045** | 1.00 | 0.87 - 1.15 | 0.993 |

[a] OR < 1.0 is associated with decreased risk and OR > 1.0 is associated with increased risk of congenital HCMV transmission. Bold indicates statistical significance ($p < 0.05$). n = 72 mother-infant dyads

**Supplementary Table 6. Univariate logistic regression analysis of antibody responses and risk of cCMV infection excluding HCMV-specific IgM+ mothers**

| Antibody variable | Maternal sera | | | Cord blood sera | | |
|---|---|---|---|---|---|---|
| | OR[a] | 95% CI | p value | OR | 95% CI | p value |
| **HCMV ADCC** | 0.89 | 0.78 - 1.01 | 0.081 | 0.86 | 0.73 - 1.01 | 0.070 |
| **HCMV IgG FcγRIII activation** | 1.07 | 0.71 - 1.07 | 0.755 | 1.08 | 0.75 - 1.57 | 0.672 |
| **UL16 IgG binding level level** | 0.92 | 0.68 - 1.23 | 0.572 | 0.87 | 0.64 - 1.18 | 0.386 |
| **UL16 IgG FcγRIII V158** | 0.78 | 0.63 - 0.96 | **0.017** | 0.72 | 0.58 - 0.90 | **0.005** |
| **UL16 IgG FcγRIII F158** | 0.80 | 0.68 - 0.95 | **0.013** | 0.76 | 0.63 - 0.92 | **0.005** |
| **UL16 IgG FcγRIII activation** | 0.86 | 0.73 - 1.01 | 0.068 | 1.01 | 0.87 - 1.17 | 0.895 |

[a] OR < 1.0 is associated with decreased risk and OR > 1.0 is associated with increased risk of congenital HCMV transmission. Bold indicates statistical significance ($p < 0.05$). n = 68 mother-infant dyads

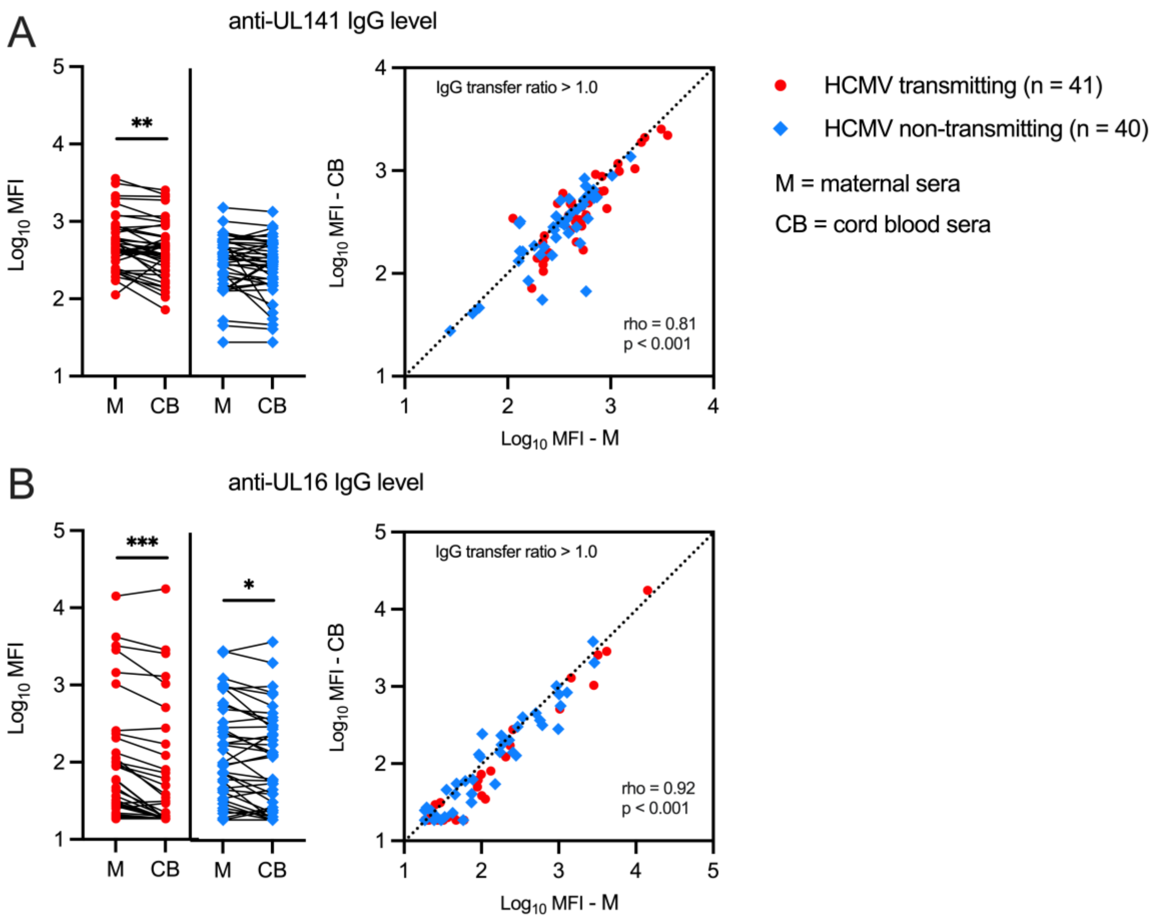


**Supplementary Figure 3. Anti-UL141 and anti-UL16 IgG transfer from maternal to cord blood sera.** Anti-UL141 and anti-UL16 IgG binding levels were measured with a binding level antibody multiplex assay using maternal (M) and cord blood (CB) sera from HCMV transmitting (red circles, n = 41) and non-transmitting (blue diamonds, n = 40) mother-infant dyads. (A) Anti-UL141 IgG level compared within mother-infant dyads and scatterplot showing Spearman correlation between anti-UL141 IgG level in paired maternal versus cord blood sera samples. (B) Anti-UL16 IgG level compared within mother-infant dyads and scatterplot showing Spearman correlation between anti-UL141 IgG level in paired maternal versus cord blood sera samples. FDR-corrected *P* values for Wilcoxon signed-rank test. * *P* < 0.05, ***P* < 0.01, ****P* < 0.001.
